# Integrative Analysis Reveals a Molecular Stratification of Systemic Autoimmune Diseases

**DOI:** 10.1101/2020.02.21.20021618

**Authors:** Guillermo Barturen, Sepideh Babaei, Francesc Català-Moll, Manuel Martínez-Bueno, Zuzanna Makowska, Jordi Martorell-Marugán, Pedro Carmona-Sáez, Daniel Toro-Domínguez, Elena Carnero-Montoro, María Teruel, Martin Kerick, Marialbert Acosta-Herrera, Lucas Le Lann, Christophe Jamin, Javier Rodríguez-Ubreva, Antonio García-Gómez, Jorge Kageyama, Anne Buttgereit, Sikander Hayat, Joerg Mueller, Ralf Lesche, Maria Hernandez-Fuentes, Maria Juarez, Tania Rowley, Ian White, Concepción Marañón, Tania Gomes Anjos, Nieves Varela, Rocío Aguilar-Quesada, Francisco Javier Garrancho, Antonio López-Berrio, Manuel Rodriguez Maresca, Héctor Navarro-Linares, Isabel Almeida, Nancy Azevedo, Mariana Brandão, Ana Campar, Raquel Faria, Fátima Farinha, António Marinho, Esmeralda Neves, Ana Tavares, Carlos Vasconcelos, Elena Trombetta, Gaia Montanelli, Barbara Vigone, Damiana Alvarez-Errico, Tianlu Li, Ricardo Blanco Alonso, Alfonso Corrales Martínez, Fernanda Genre, Raquel López Mejías, Miguel A. Gonzalez-Gay, Sara Remuzgo, Begoña Ubilla Garcia, Ricard Cervera, Gerard Espinosa, Ignasi Rodríguez-Pintó, Ellen De Langhe, Jonathan Cremer, Rik Lories, Doreen Belz, Nicolas Hunzelmann, Niklas Baerlecken, Katja Kniesch, Torsten Witte, Michaela Lehner, Georg Stummvoll, Michael Zauner, Maria Angeles Aguirre-Zamorano, Nuria Barbarroja, Maria Carmen Castro-Villegas, Eduardo Collantes-Estevez, Enrique de Ramon, Isabel Díaz Quintero, Alejandro Escudero-Contreras, María Concepción Fernández Roldán, Yolanda Jiménez Gómez, Inmaculada Jiménez Moleón, Rosario Lopez-Pedrera, Rafaela Ortega-Castro, Norberto Ortego, Enrique Raya, Carolina Artusi, Maria Gerosa, Pier Luigi Meroni, Tommaso Schioppo, Aurélie De Groof, Julie Ducreux, Bernard Lauwerys, Anne-Lise Maudoux, Divi Cornec, Valérie Devauchelle-Pensec, Sandrine Jousse-Joulin, Pierre-Emmanuel Jouve, Bénédicte Rouvière, Alain Saraux, Quentin Simon, Montserrat Alvarez, Carlo Chizzolini, Aleksandra Dufour, Donatienne Wynar, Attila Balog, Márta Bocskai, Magdolna Deák, Sonja Dulic, Gabriella Kádár, László Kovács, Qingyu Cheng, Velia Gerl, Falk Hiepe, Laleh Khodadadi, Silvia Thiel, Emanuele de Rinaldis, Sambasiva Rao, Robert J.Benschop, Chris Chamberlain, Ernst R. Dow, Yiannis Ioannou, Laurence Laigle, Jacqueline Marovac, Jerome Wojcik, Yves Renaudineau, Maria Orietta Borghi, Johan Frostegård, Javier Martín, Lorenzo Beretta, Esteban Ballestar, Fiona McDonald, Jacques-Olivier Pers, Marta E. Alarcón-Riquelme

**Affiliations:** Department of Medical Genomics, Center for Genomics and Oncological Research (GENYO), Granada, Spain; Pharmaceuticals Division, Bayer Pharma Aktiengesellschaft, Berlin, Germany; Cancer Epigenetics and Biology Program, Bellvitge Biomedical Research Institute (IDIBELL), Barcelona, Spain; Department of Bioinformatics, Center for Genomics and Oncological Research (GENYO), Granada, Spain; Institute of Parasitology and Biomedicine López Neyra, Spanish National Research Council, Granada, Spain; Université de Brest, INSERM, Labex IGO, CHU de Brest, Brest, France; UCB Pharma, Slough, United Kingdom; Andalusian Public Health System Biobank, Granada, Spain; Unidade de Imunologia Clínica, Centro Hospitalar do Porto, Portugal; Serviço de Imunologia EX-CICAP, Centro Hospitalar e Universitário do Porto, Porto, Portugal; Laboratorio di Analisi Chimico Cliniche e Microbiologia - Servizio di Citofluorimetria, Fondazione IRCCS Ca’ Granda Ospedale Maggiore Policlinico di Milano, Milano, Italy; Scleroderma Unit, Referral Center for Systemic Autoimmune Diseases, Fondazione IRCCS Ca’Granda Ospedale Maggiore Policlinico di Milano, Milan, Italy; Servicio Cántabro de Salud, Hospital Universitario Marqués de Valdecilla, IDIVAL, Santander, Spain; Hospital Clinic, Institut d’Investigacions Biomèdiques August Pi i Sunyer, Barcelona, Catalonia, Spain; Skeletal Biology and Engineering Research Center, KU Leuven and Division of Rheumatology, UZ Leuven; Klinikum der Universitaet zu Koeln, Cologne, Germany; Klinik für Immunologie und Rheumatologie, Medical University Hannover, Hannover, Germany; Medical University Vienna, Vienna, Austria; Rheumatology Service, Reina Sofia Hospital, Maimonides Institute for Research in Biomedicine of Cordoba (IMIBIC), University of Cordoba, Cordoba, Spain; Servicio Andaluz de Salud, Hospital Regional Universitario de Málaga, Spain; Servicio Andaluz de Salud, Complejo Hospitalario Universitario de Granada (Hospital Universitario San Cecilio), Spain; Servicio Andaluz de Salud, Complejo Hospitalario Universitario de Granada (Hospital Virgen de las Nieves), Spain; Università degli studi di Milano, Milan, Italy; Istituto Auxologico Italiano, Milan, Italy; Pôle de Pathologies Rhumatismales Inflammatoires et Systémiques, Institut de Recherche Expérimentale et Clinique, Université Catholique de Louvain, Brussels, Belgium; AltraBio SAS, Lyon, France; Immunology & Allergy, University Hospital and School of Medicine, Geneva, Switzerland; University of Szeged, Szeged, Hungary; Department of Rheumatology and Clinical Immunology, Charité University Hospital, Berlin, Germany; Precision Immunology Cluster, Sanofi, Cambridge, MA, USA; Sanofi Genzyme, Framingham, MA, USA; Eli Lilly and Company, Indianapolis, IN, United States of America; Institut de Recherches Internationales Servier, Suresnes, France; QuartzBIO, SA, Geneva, Switzerland; Unit for Chronic Inflammatory Diseases, Institute for Environmental Medicine, Karolinska Institutet, Stockholm, Sweden

**Keywords:** autoimmunity, systemic lupus erythematosus, rheumatoid arthritis, systemic sclerosis, primary Sjogren’s syndrome, mixed connective tissue disease, primary antiphospholipid syndrome, undifferentiated connective tissue disease, SNF, stratification, classification criteria

## Abstract

**Background:** Clinical heterogeneity, a hallmark of systemic autoimmune diseases (SADs) impedes early diagnosis and effective treatment, issues that may be addressed if patients could be grouped into a molecular defined stratification.

**Methods:** With the aim of reclassifying SADs independently of the clinical diagnoses, unsupervised clustering of integrated whole blood transcriptome and methylome cross-sectional data of 918 patients with 7 SADs and 263 healthy controls was undertaken. In addition, an inception cohort was prospectively followed for 6 and 14 months to validate the results and analyze if cluster assignment changed or not with time.

**Results:** Four clusters were identified. Three clusters were aberrant, representing ‘inflammatory’, ‘lymphoid’, and ‘interferon’ patterns each including all diagnoses and defined by genetic, clinical, serological and cellular features. A fourth cluster showed no specific molecular pattern and accumulated also healthy controls. An independent inception cohort showed that with time, the molecular clusters remain stable, showing that single aberrant molecular signatures characterize each individual patient.

**Conclusions:** Patients with SADs can be jointly stratified into three stable disease clusters with specific molecular patterns differentiating different molecular disease mechanisms. These results have important implications for future clinical trials and the study of therapy non-responsiveness marking a paradigm shift in the view of SADs.

## INTRODUCTION

The systemic autoimmune diseases (SADs) are entities diagnosed based on different clinical and laboratory criteria. The diseases are highly heterogeneous with varied progression of disease severity. In general, the time from disease onset to diagnosis can be of many years, leading to damage accrual and poor prognosis. Moreover, some individuals never fulfill the clinical criteria for a specific SAD and remain undiagnosed for years or a lifetime (undifferentiated connective tissue disease, UCTD).

It is known that SAD patients from different diagnoses share clinical features. A number of patients with systemic lupus erythematosus (SLE) may develop joint deformities in hands and feet, similar to those found in rheumatoid arthritis (RA), albeit without erosions, and all may share autoantibody specificities^1^. Mixed connective tissue disease (MCTD) patients may have clinical manifestations usually observed in SLE, RA or systemic sclerosis (SSc)^2,3^. While patients with SLE and RA may present with secondary Sjögren’s syndrome (SjS), many patients have the primary entity (pSjS), without evidence of RA or SLE^4^. Similarly, SLE patients may have secondary antiphospholipid syndrome, but there are patients with primary antiphospholipid syndrome (PAPS) who tend not to develop SLE, even after many years follow-up^5^. This overlapping clinical landscape hinders diagnosis and the early treatment.

Genetic studies have shown that SADs share susceptibility genes^6^ and molecular features, such as increased expression of interferon inducible genes (interferon signature)^7,8^, mainly observed in SLE patients. But not all patients with SLE have the interferon signature. Some patients with SSc have disease limited to the skin^9^, and not all patients who fulfill the diagnostic criteria for RA have anti-citrullinated peptide antibodies (∼70%)^10^. A number of patients with SLE and pSjS share the presence of anti-SSA and anti-SSB antibodies, and in this regard, also share alleles of the HLA class II gene DRB1*0301^11^. Thus, this heterogeneity impedes the identification of new therapies and has consequences for the selection of response endpoints and the overall results of clinical trials hindering the advance of new medications^12,13^. Therefore, development of new therapies, prescription of existing ones, and even the early diagnosis of SADs might benefit from a uniform molecular classification that allows their stratification and considers their commonalities.

Some efforts have been made in order to stratify individual SADs into homogeneous molecular groups of patients^12-14^, and very recently, to reclassify three different autoimmune clinical outcomes into a molecular classification based solely on mass spectrometry^15^. All these studies support the hypothesis that molecular reclassification is feasible, but they lack the sufficient number of patients, and multiple layers of information needed for this purpose and its validation. Thus, in an unprecedented study in systemic autoimmunity, high dimensional molecular data from whole blood shows how seven SADs (SLE, RA, SSC, SjS, MCTD, PAPS, and UCTD) stratify into groups of molecular patterns that are stable over time, each having defined serological, genetic and clinical characteristics.

## RESULTS

### Integrative molecular analysis redefines the distribution of systemic autoimmune diseases into functional clusters that are independent of the diagnoses

Genome wide transcriptome and methylome information from a discovery set of 722 patients with SADs was used in an unsupervised protocol to perform an integrative molecular analysis. This approach unveiled 4 clusters of patients, characterized by WGCNA^16^ functional modules comprising 5 modules of elevated gene expression, and 3 modules of CpG hypomethylation (Figure S3). The same results were observed in an independent validation set of 196 patients with SADs. These modules formed specific molecular signatures that defined the clusters (Figure 1).

**Figure 1.**
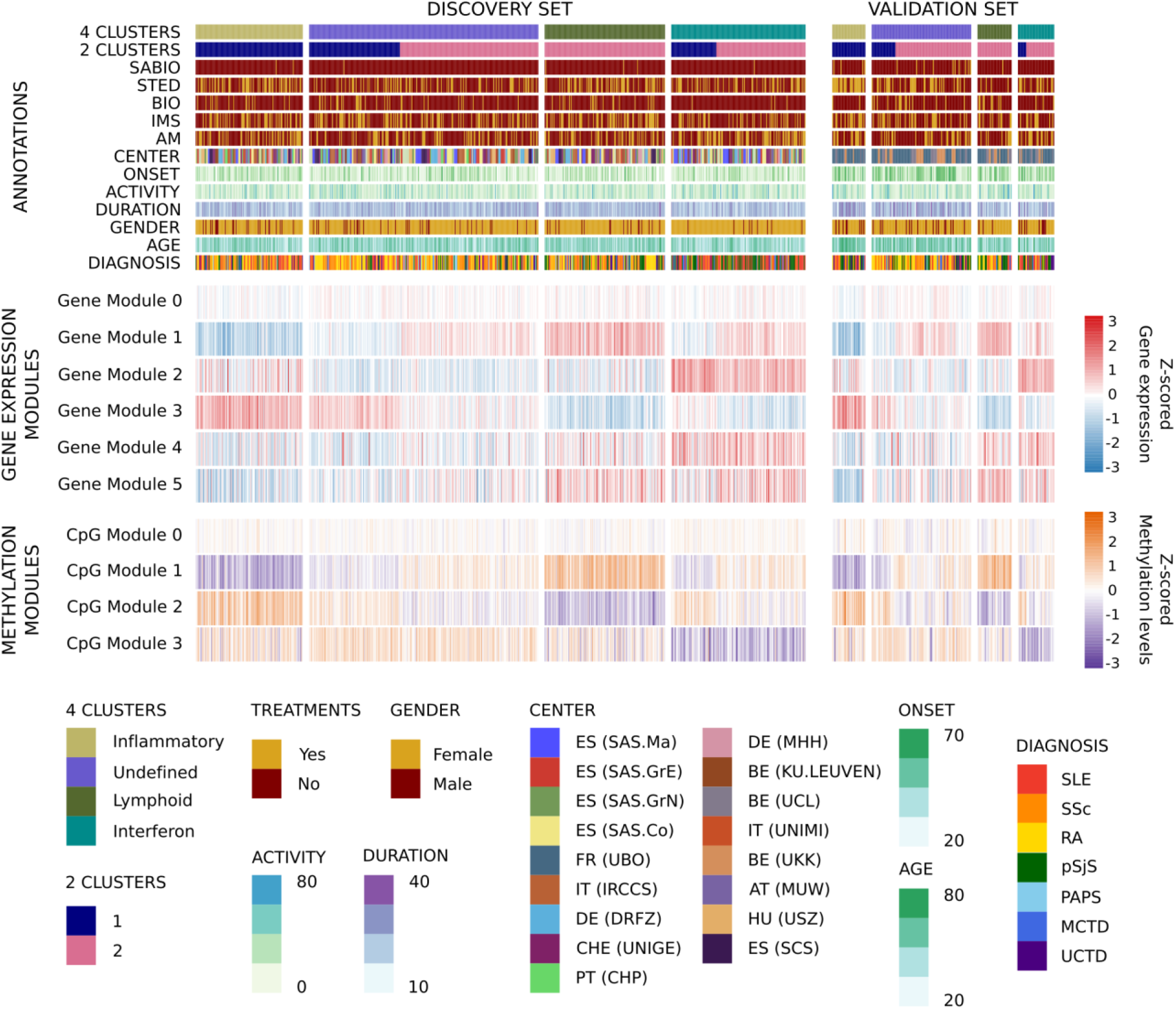
The molecular pattern distribution of SADS is limited to 4 clusters. Heatmap showing the distribution of gene and CpG functional modules across the 4 autoimmune disease clusters. In columns patients are grouped by cluster assignment and in rows the functional modules of the features are shown with their scaled median values. The two subsets of patients comprising the discovery and validation sets are shown. For the transcriptome, red represents over-expression and blue represents under-expression. For the methylome, purple represents hypo-methylation and orange represents hyper-methylation. At the top of the figure the annotation shows: two configurations of clusters for 4 (4 CLUSTERS) and 2 (2 CLUSTERS) groups, each of the treatment groups for each individual (SABIO, systemic antibiotics; STED, steroids; BIO, biologics; IMS, immunosuppressors and AM, antimalarials), recruitment centers distribution, disease activity as physician global assessment, duration of the disease since diagnosis, age at onset, gender, age and diagnosis. The abbreviations for the recruitment centers can be found in Table S1.

Importantly, gene and CpG modules showed high functional concordance according to Chaussabel et al.^17^ and Li et al.^18^ dataset enrichments. Overexpressed gene modules and hypomethylated CpG modules in the same clusters were enriched with the same functionalities (Figure 2A). First, an inflammatory cluster was defined by overexpression and hypomethylation of genes and CpGs included in modules driven by monocytes and neutrophils (gene module 3 and CpG module 1). A lymphoid cluster was composed of T and natural killer (NK) cell functions (gene module 1 and CpG module 2), while an interferon cluster was defined by interferon and viral and dendritic cell functions (gene module 2 and CpG module 3). One cluster had no clear defined functional modules (undefined cluster). Other functionalities complemented the molecular information. Cell cycle and transcription upregulation (gene module 4) was associated with the interferon cluster, and B lymphocyte functions (gene module 5) were particularly observed in both the lymphoid and interferon clusters (Figure 2A).

**Figure 2.**
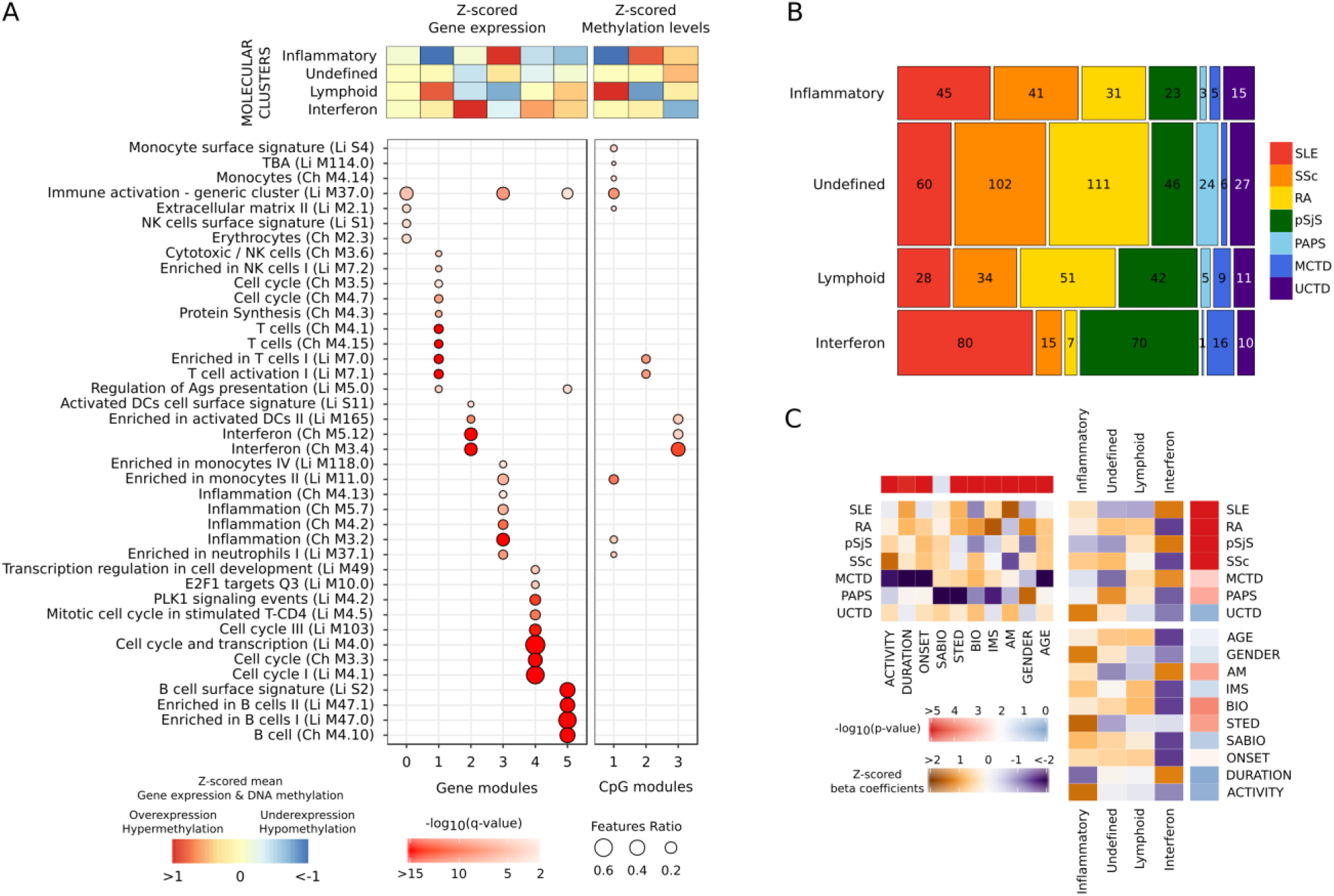
OMIC layers of information functionally characterize each of the molecular clusters, do not follow the clinical diagnoses, and are not conditioned by confounders. (A) Annotation of the selected features according to the hypergeometric enrichment of their modular functional assignment. The module annotations were obtained using the blood immunological signature databases of Chaussabel et al. (Ch) and Li et al (Li). Only significant enrichment results are shown (q-value < 0.01). Significant modules are shown in columns and their annotation in rows. (B) Mosaic plot showing the distribution of diseases in each cluster with the number of individuals inside each block. Diseases are represented by a color in columns. (C) Association of covariates with clinical diagnosis, molecular clusters, and the associations between them. The significance of the associations is represented as a scale ranging from blue (non-significant) to red (significant). The direction of the association is shown as the z-scored beta coefficients where orange is enrichment and purple is depletion.

Patients with all the clinical diagnoses could be found in all 4 clusters (Figure 2B). The interferon and the undefined clusters presented an enrichment of some diagnoses (Figure 2C). The undefined cluster had an increased number of RA, SSc and PAPS, and grouped around 40% of all patients. The interferon cluster was enriched in SLE, pSjS, and MCTD (Figure 2B). The inflammatory and lymphoid clusters had no particular enrichment. Interestingly, UCTD patients were the only ones not significantly enriched or depleted in any cluster (Figure 2C) but were evenly distributed across all clusters. Most MCTD diagnosis, whose existence as a disease entity has been controversial^3,19^,20 fell in the interferon cluster enriched in SLE and pSjS.

Except for systemic antibiotics, covariates associated with the transcriptome and the methylome principal components (Figure S1) were unevenly distributed across the clinical diagnoses (Figure 2C). Importantly, most of these covariates did not show a dependency with the molecular clusters (Figure 2C). The significant association that remained after adjustment of some treatments with the clusters (antimalarials, biologicals and steroids) was due to the enrichment of the clinical diagnoses (Figure 2C) and not because of the treatments themselves. For example, hydroxychloroquine treated patients were enriched in the interferon cluster, and this association was driven by the enrichment of SLE, pSjS and MCTD (Table S4). Importantly, time since diagnosis was not significantly enriched (Duration in Figure 2C).

In addition, associations between the CpGs and genes in the functional modules revealed different regulatory relationships. *Cis* associations linked CpG modules with their counterpart gene modules, while *trans* associations did not show major relationships between homologous functional modules (Figure S4A). A major difference between clusters was that more than 80% of the CpGs in the interferon modules were associated in *cis* with genes in the interferon gene modules, whereas most of the features in the rest of the modules had few (11 to 17%) *cis* associations (Figure S4B). Thus, these complex relationships between methylation and gene expression modules revealed a deeper view of the molecular state of the patients than what a single layer, say gene expression, may give.

Analyzing a deeper layer, cell-type specific histone marks and transcription factor binding site (TFBSs) analysis (Figures S4C and S4D) confirmed their functional relationship with the modules (Supplementary Information).

### Low dimensional layers of information show specific patterns in the molecular stratification

Data on pre-selected autoantibodies, cytokines, small lipid moiety (natural) autoantibodies and cell surface antigen markers^21^ were used to characterize the clusters (Supplementary Information). The serology characterization showed four patterns of associations with the clusters: anti-citrullinated peptide, anti-centromere B and IgM anti-phosphatidylcholine natural autoantibodies were slightly enriched in the lymphoid and the undefined clusters, and a strong depletion was observed in the interferon cluster; the interferon cluster showed an enrichment of anti-dsDNA, anti-Sm, anti-SSA, anti-SSB, anti-U1RNP and protein free light chains, and an increase in IP-10, BAFF, MCP-2 and TNF-α; the inflammatory cluster was increased in MMP-8 and C-reactive protein; while high levels of IL-1RA and BLC were shared by the inflammatory and interferon clusters (Figure 3A). In general, the association of serological markers followed the molecular functions that defined the clusters. For example, the interferon cluster was associated with cytokines regulated by type I IFNs, such as IP-10 and BAFF, but also with TNFα, which, under some situations, was shown to induce type I interferon^22^. On the other hand, patients from the inflammatory cluster showed elevated levels of C-reactive protein and MMP-8, both markers of acute inflammatory processes^23,24^.

**Figure 3.**
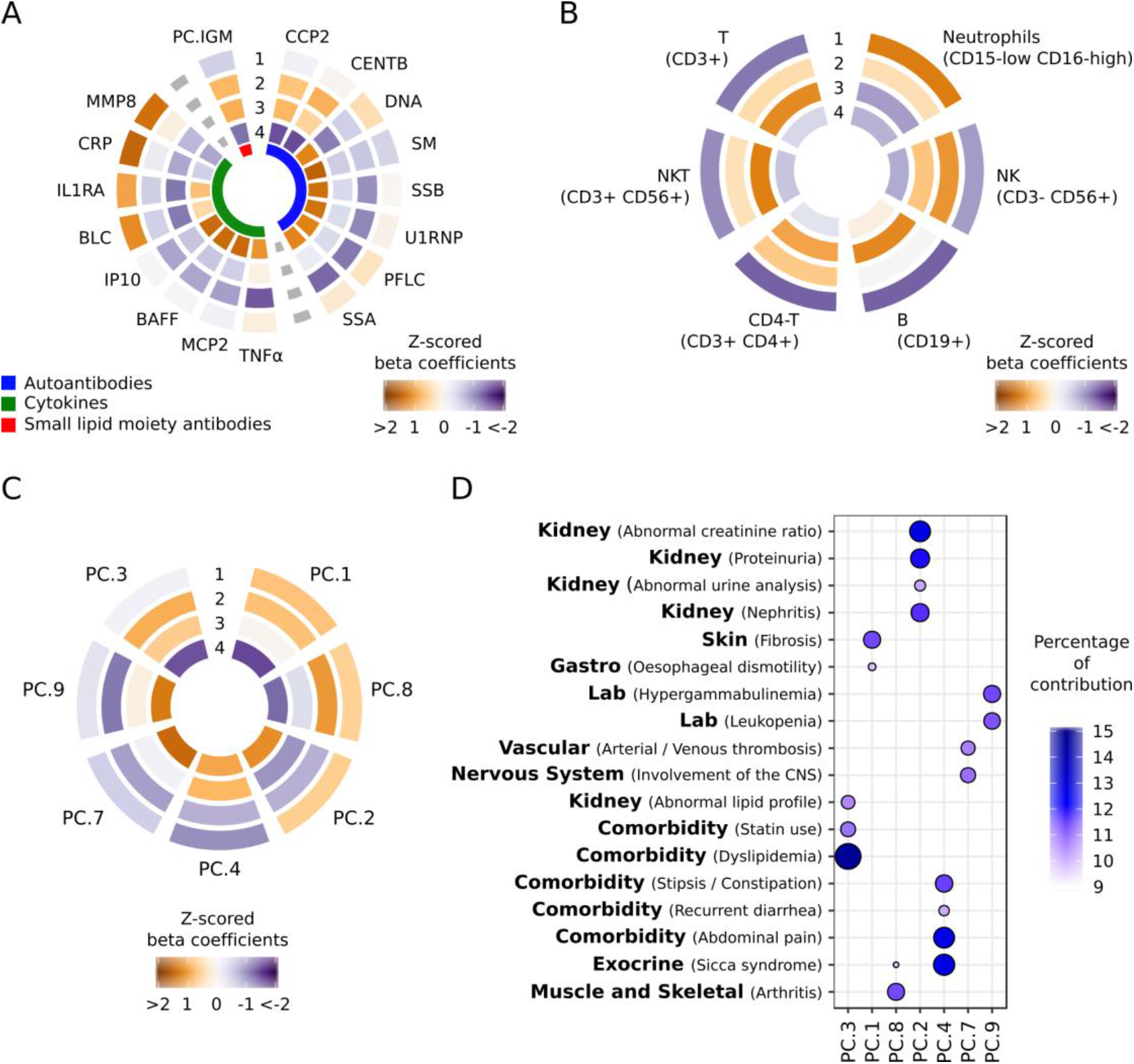
Each cluster is associated with specific low-dimensional layers of clinical information. Z-scored beta coefficients of the significant ANOVA associations (FDR < 0.01) for (A) serology, (B) flow-cytometry, and (C) clinical data, are shown. The serology information included autoantibodies (blue), cytokines (green), and antibodies against small lipid moieties or natural autoantibodies (red). Abbreviations are as follows: CCP2, anti-citrullinated peptide antibodies; CENTB, anti-centromere B; DNA, anti-dsDNA; SM, anti-Sm; SSB, anti-SSB or anti-La; U1RNP, anti-U1RNP; PFLC, Protein free light chains; SSA, anti-SSA or anti-Ro; PC.IGM, IgM anti-phosphatydilcholine. Hierarchical clustering of the results expressed as Z-scored beta coefficients are represented in the circle plots (A, B, and C). The four autoimmune molecular clusters are shown concentrically from the outermost circle with cluster 1 (inflammatory), followed by 2 (undefined), 3 (lymphoid), and 4 (interferon), in the innermost circle. Clinical information was summarized using principal components (PC) and the PCs most significantly associated with each cluster are shown. (D) Only those clinical items having a significant contribution to each significant PC (observed contribution higher than five times the expected) are depicted. Features and PCs are sorted by hierarchical clustering.

The cell population composition of the clusters revealed a high proportion of neutrophils in the inflammatory cluster, a slightly high proportion of NK cells in the undefined cluster, T cells, B cells, NK cells and NKT cells in lymphoid cluster, and with the exception of a slight B cell increase, the interferon cluster did not show any particular cellular enrichment (Figure 3B). In agreement with other data, this result reflects the expression of an interferon signature by all cells^25^.

The genetic contribution of risk alleles known for the four main SADs was analyzed on the clusters (Figure S5D and S5E). The interferon cluster was particularly enriched in HLA (Human Leukocyte Antigen) class II risk alleles, but no other cluster showed significant enrichment. In order to confirm this result, four GWAS were performed comparing each cluster and healthy controls. Again, the only signal with genome wide significance level (*p-value* < 5e10^−8^) came from alleles located in HLA class II genes within the region covering *HLA-DRA, DRB5, DRB1, DQA1, DQB1, DQA2, DQB2*, and *DOB*) in the interferon cluster (Figure S6). The weak HLA association found in the other clusters was located within the HLA class I gene region (Supplementary Files). This result implies that the genetic associations observed for some of the systemic autoimmune diseases (i.e. SLE) actually reflect the molecular mechanisms occurring in those individuals whose molecular disease pathway is the type I interferon and not any of the other.

The clinical information of the patients was summarized into principal components and associations were found for each molecular cluster (Figure 3C and 3D). The interferon cluster was associated with some of the most extreme phenotypes in SADs such as kidney function abnormalities (including nephritis), thrombosis, nervous system involvement, and leukopenia, in addition to other minor comorbidities. Fibrosis complications in both skin and muscle skeletal organs were enriched in the inflammatory cluster and in the undefined cluster, while kidney clinical feature associations were enriched in the inflammatory cluster and not in the undefined cluster. Comorbidities related with abnormal lipid metabolism and dyslipidemias were enriched in the undefined cluster. The lymphoid cluster, in general, presented less aggressive phenotypes than the other clusters, with enrichment of dyslipidemias, presence of abdominal pain, diarrhea, and constipation. Association with *sicca* syndrome was also found in this cluster.

### Undefined cluster shows a healthy-like molecular pattern

To gain insight on the type of patients that were being grouped in the undefined cluster, different analyses were performed. Healthy individuals were assigned to the molecular clusters by means of the clustering model obtained with the discovery set. Of these, 74% grouped in the undefined cluster, compared to 12%, 11% and 3% that were assigned to the inflammatory, lymphoid and interferon clusters, respectively (Figure 4A). Additionally, differential expression analysis was performed between each patient cluster and healthy controls (Figure 4D). The highest number of differentially expressed genes (DEGs) was found for the inflammatory cluster (2898 DEGs), followed by the interferon cluster (820 DEGs) and the lymphoid cluster (294 DEGs), while only 9 genes were differentially expressed in the undefined cluster between patients and controls. The low amount of DEGs in this cluster suggested that it could be due to two non-mutually exclusive reasons. Some of the most enriched diseases in this cluster, RA and SSc, could be undergoing processes in the target tissues (synovia and skin, respectively), and blood was unable to identify an aberrant molecular pattern. On the other hand, this undefined cluster might also be grouping patients undergoing remission or having low disease activity, related to the inclusion criteria used to recruit the patients. To test this, disease activity was compared between clusters. Disease activity indexes are designed for each clinical diagnosis through different scores measuring specific clinical manifestations^26^, and are not performed across all diseases. For this analysis 138 SLEDAI-scored, 79 ESSDAI-scored and 17 DAS28-scored SLE, SjS and RA patients, respectively, were available. In both, SLEDAI and ESSDAI, higher disease activity scores were shown in all clusters as compared to the undefined cluster (Figure 4B and 4C). Significant differences were found for the inflammatory and interferon clusters when compared with the undefined cluster in the SLEDAI analysis (p=0.04 and p=0.02, respectively, Wilcoxon rank sum test), while the ESSDAI analysis showed suggestive non-significant p-values. The results suggest that low disease activity could lie behind the undifferentiated patterns of this cluster. Thus, given these results the undefined cluster observed might be considered to have a healthy-like molecular pattern.

**Figure 4.**
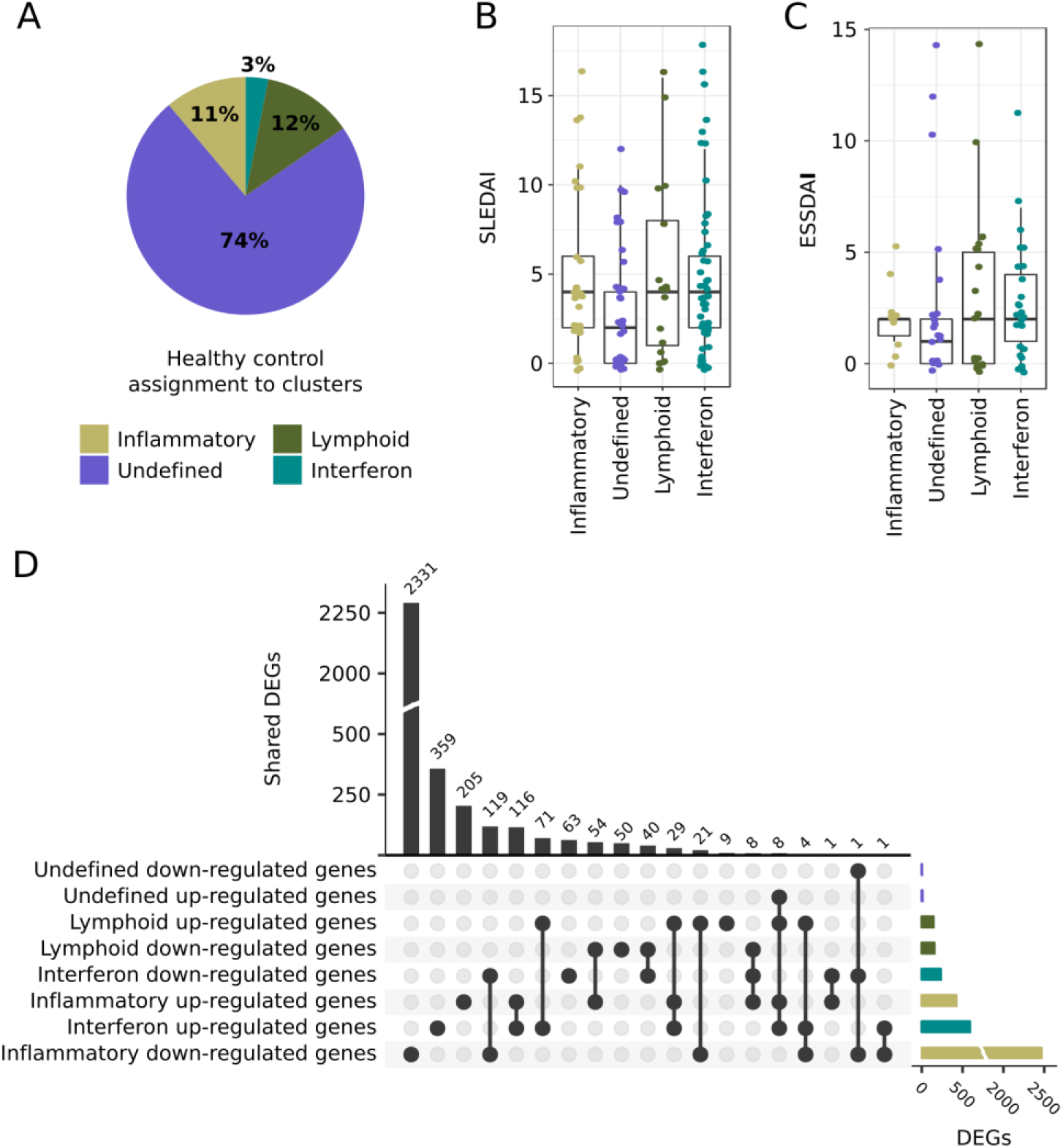
Three of the clusters present aberrant molecular patterns that may be reverted by specific drugs. (A) Distribution of healthy individual assignments to the SADs molecular classification. The pie diagram shows that nearly 74% of controls are similar to patients in the undefined molecular cluster. (B) SLEDAI boxplots by molecular cluster, n=138 SLE patients. (C) ESSDAI boxplots by molecular cluster, n=79 SjS patients. In both boxplots, the lower and upper hinges correspond to the first and third quartiles, while whiskers represent the 1.5 * interquartile ranges. (D) Differentially expressed genes (DEGs) between clusters and healthy controls. Top bar plot (black bars) shows shared DEGs across clusters, right bar plot (colored bars) represents the number of DEGs by cluster. The dot connectivity plot represents the intersections across clusters.

### Pathological molecular patterns are stable over time

It is possible that the clusters represent the disease state of the individual patients at a given point in time and that patients could “move”, with time, to different clusters as disease progresses. Furthermore, the long time of disease combined with years of treatment (12 years on average since diagnosis for the CS cohort) could lie behind the configuration of clusters that we observe. In order to determine whether the clusters could be observed in patients with newly diagnosed disease and if these were stable over time, an independent and newly recruited inception cohort was analyzed using the model developed using the discovery set. Furthermore, the inception cohort could also be a third validation set.

Indeed, first, the clusters observed were exactly the same as those identified with the discovery set and observed again in the validation set, confirming once more the clustering model. Second, the stability values for patients with information at recruitment and at 6 (n=103) or 14 months (n=78) samplings showed similar results (Figure 5A and 5B). In both comparisons, most of the patients remained in the same cluster after recruitment (62-63%) and 34-33% of them moved from a pathological cluster (inflammatory, lymphoid or interferon cluster) to the healthy-like cluster or vice versa, while only 4% of patients “moved” to a different pathological cluster. The analysis of all three time-points together (n=68) showed that only 4 patients (6%) changed between different pathological clusters, while 33 patients (48%) remained in the same cluster over all three-time points (Figure 5C). The remaining patients (46%) showed a behavior that resembled a relapse-remission dynamic of SADs from a molecular point of view: their pathological cluster was stable (never assigned to a different pathological cluster) but they could be assigned at a given time-point to the healthy-like cluster (Figure 5C).

**Figure 5.**
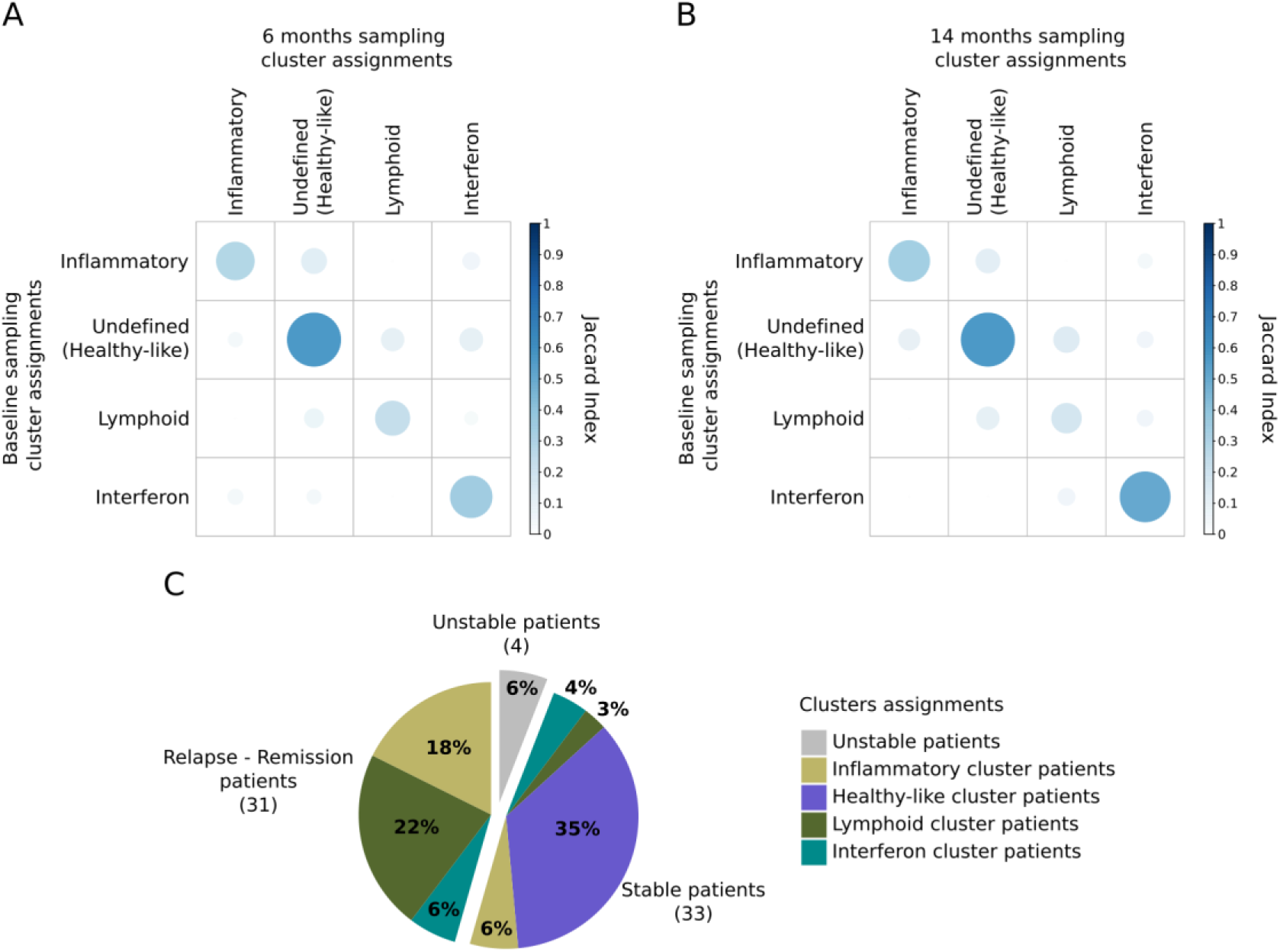
Aberrant molecular patterns are stable in time and related to relapsing moments of disease. (A) The stability Jaccard index between molecular cluster assignments at baseline (rows) and at 6 months (columns). (B) Stability Jaccard index between molecular cluster assignments at baseline (rows) and at 14 months (columns). (C) Patient classification considering the 3 time point cluster assignments. The definitions are as follows: stable patients, patients assigned to exactly the same cluster in the 3 time points; relapse-remission patients, patients assigned to only one pathological cluster but have been at any given time point in the healthy-like cluster; unstable patients, patients assigned to more than one aberrant cluster at any one time point.

## DISCUSSION

With an unsupervised model integrating transcriptome and methylome data we observe 3 aberrant and pathological clusters that we described as an inflammatory cluster with a pattern coming from neutrophils and monocytes, a lymphoid cluster enriched in lymphocytes, particularly T cells, NK cells, NKT cells but also B cells, and an interferon cluster spread across all cell types and with a slight enrichment of B cells. Additionally, an undefined molecular cluster was found, where most healthy controls were assigned and indeed with very few differentially expressed genes as compared to controls. Genetically, only the interferon cluster has a significant association with the HLA class II locus. This locus is a well-known ubiquitous risk factor across SADs, but only the interferon cluster presented this genetic association. This significant association reflects two things: on the one hand that the HLA-DRB1 is clearly associated with a subgroup of SLE and pSjS (and MCTD), secondly, having such a strong association in a relatively small number of patients reflects the reliability of the classification method, grouping patients from different SADs that share common pathological molecular patterns that might be driven by a common genetic background. Additional genetic analysis in larger groups of patients using our classification approach, might lead to the discovery of additional genes that have currently been associated with different clinical outcomes.

Our results from an independent inception cohort suggest that patients’ cluster assignment is independent of the time since diagnosis, not dependent on treatment and stable over time. This implies that each patient has a single mechanistic profile of disease pathology out of 3 possible, when detectable through the integrated transcriptome and methylome obtained from blood. The predominance of patients assigned to the healthy-like cluster in the inception cohort reflects the decision taken during the design of the study that patients would not be heavily treated at baseline in order to analyze if therapy given afterwards would interfere with the results. Therefore, this resulted in a large number of patients that remained with low disease activity and hence in the healthy-like cluster. Nevertheless, patients were treated as required in the follow-up, supporting that therapy did not condition the structure of the clusters.

The molecular analyses of this study were performed on whole blood, which is not the final target tissue of most diseases. Fortunately, most of the cell-types implicated in autoimmune pathologies are infiltrated from blood to tissues. Thus, some of the aberrant signatures in the tissues can also be detected in blood^27,28^ but of course not all of them^29^. On the other hand, having now an algorithm and a model that defines the structure in blood of the patients, interventional clinical trials can use the modelling method with much fewer patients (even just one) to assign these to the clusters and follow disease progression in relation to the drugs to be tested. Therefore, we open the door to have a closer look and identify endophenotypes or pathotypes that relate to these blood clusters. Future for single cell studies may distinguish cell-specific and tissue-specific mechanisms as a sub-stratification of patients considering the blood cluster to which a patient belongs. For example, we observe that kidney disease is enriched in the inflammatory and interferon clusters, leading to the question whether the kidney disease of the patients in each cluster may show different pathotypes.

This study shows for the first time and in an unprecedented number of individuals that systemic autoimmune disease patients with 7 different clinical diagnoses share molecular clusters defined by specific molecular patterns stable over time. The clusters have specific clinical and serological characteristics, but also quite different regulatory landscapes and suggest that three alternative and different molecular pathways drive the disease in each individual. Further, only the interferon cluster had an important genetic component with a very strong association with the HLA class II risk alleles. The results obtained in this study are a first step towards laying the foundations for the personalized medicine in systemic autoimmune diseases.

## METHODS

### Samples and Data types

Two cohorts of individuals with 7 different SADs were recruited: a cross sectional cohort composed of 2003 patients and 617 healthy controls, and an inception cohort of 215 patients followed and sampled at 0, 6 and 14 months. Inclusion and exclusion criteria are detailed in Table S2. Demographic information and treatment are given in Tables S3-S6. A detailed description of both cohorts can be found in the Supplementary Information.

Blood and serum samples were obtained from all patients. High-dimensional genome wide genotype, transcriptome, DNA methylome and proportions of relevant cell types were analyzed from whole blood samples. Low dimensional information was obtained from serum samples, including selected serology information such as SADs related autoantibodies, cytokines, chemokines and inflammatory mediators. A detailed description of all the protocols and methods can be found in the Supplementary Information.

The Ethical Review Boards of the 18 participating institutions approved the protocol of the cross sectional study. In addition, the boards of the 6 sites involved approved the inception study protocol. The studies adhered to the standards set by International Conference on Harmonization and Good Clinical Practice (ICH-GCP), and to the ethical principles that have their origin in the Declaration of Helsinki (2013). The protection of the confidentiality of records that could identify the included subjects is ensured as defined by the EU Directive 2001/20/EC and the applicable national and international requirements relating to data protection in each participating country. The CS study is registered with number NCT02890121, and the inception study with number NCT02890134 in ClinicalTrials.gov.

### High-dimensional datasets quality control

Each high-dimensional OMIC dataset (genomics, transcriptomics and methylomics) was quality controlled in order to discard samples and features with low quality due to technical issues, and only individuals considered Europeans in the demographic record were kept. After platform-specific data quality controls were performed (detailed information is shown in Supplementary Information), samples with information for all OMIC datasets were selected, and shared information across platforms was compared. Briefly, gender and genotype concordance for each individual was tested using a concordance QC method combining genotype, methylation and transcriptome data (a detailed summary of the entire quality control process can be found in Table S10 and Table S12).

### Unsupervised clustering analysis

The quality-controlled dataset of patients with SADs was split into a discovery set (∼80% cases and healthy controls) and a validation set (∼20% cases). The validation set was composed of all patients from three independent recruitment centers (France, UBO; Belgium, UCL and Germany, UKK). The discovery set was corrected for batch and potential confounder effects (gender, age, treatment effects and center of recruitment biases) using linear regression models but preserving the differences between clinical conditions using the *removeBatchEffect* from the *limma R package*^30^. Annotations associated (linear regression p-value ≤ 0.05) with any of the transcriptome and/or methylome first ten principal components were considered confounders and included in the models (Figure S1). Identification analysis of the confounders was performed using the *swamp R package*^31^. After correction, samples that were identified as outliers either for transcriptome or methylome were removed from the analysis. Outliers were assessed by multidimensional scaling and a fixed threshold based on mahalanobis distance cut-off of 8 for both transcriptome and methylome (*MASS R package*^32^). In total, from the discovery cohort, 44 samples were discarded (28 due to transcriptome, and 16 samples for methylome). Outliers were not associated with any clinical feature or meta-information (data not shown).

In cancer reclassification studies, features with the highest variability across cancer types are chosen for reclassification purposes in an unsupervised way^33^, but the natural variability of the immune-system makes this approach inappropriate for systemic autoimmune diseases because of the dynamics of the immune system and its dependence on natural variability^34^. Then only features with statistically increased variability in cases (i.e. those individuals diagnosed with any SAD) compared to healthy controls were chosen (Genes and CpGs with FDR Levene-test below 0.1 and a case-versus-control variability fold-change higher than 0.5 were selected). Gene expression and DNA methylation variability contributes to disease susceptibility^35,36^, which implies that our feature selection might be able to select features which discriminate between differential molecular patterns, and at the same time remove high variable features not related with the pathologies included in the study. The feature selection procedure resulted in 1821 genes and 4144 CpGs (Figures S2A and S2B).

Similarity Network Fusion (SNF)^37^ was selected to obtain the clusters of SAD patients. Transcriptome and methylome selected features were integrated using SNF and the clustering model was trained on the discovery set by means of a 10 times nested cross-validation approach (Figure S2C). Training and testing of the clustering model were done as follows. Ten times nested cross-validation was run on the discovery set (without healthy controls) in order to set up the optimal hyperparameters for the SNF algorithm. An inner 5-fold cross-validation was performed with each training subset of the outer 10-fold cross-validation (Figure S2C). Combinations of the recommended SNF hyperparameters values were tested: 10 to 30 neighbors in the K-nearest neighbors (K) with a step of 5, 0.3 to 0.8 hyperparameter used in constructing similarity network with a step of 0.1 and 2 to 20 numbers of clusters. The hyperparameters were optimized through maximizing the proportion of cluster assignments that agree from the test datasets using the trained SNF model with the train dataset, and a model trained using each dataset independently. The average proportion of labels agreement for each combination of hyperparameters were compared with the results of randomized matrixes (random background structure), the results are shown in Figure S2D. This clustering approach showed two optimal configurations of 2 and 4 clusters of patients. The two potential cluster configurations had similar stability values (Figures S2D and S2E). However, 4 clusters configuration presented a better stability and allowed to characterize the patients deeper, and therefore was selected for further analysis. The clustering results obtained from the discovery set were confirmed using an independent validation set (Figure S2C), validation samples were assigned to each cluster by means of *groupPredict* SNF function using the model trained with the discovery set.

### Characterization of molecular signatures that drive the clusters

In order to assess the molecular signatures that drive the clusters and their relationships across layers of information, selected genes and CpGs were independently grouped into functional modules using weighted correlation network analysis, WGCNA^16^. For genes and CpGs selected and included in the clusters, meta-features (groups of correlated features) were identified using default parameters and “signed hybrid” network type^16^. Two immunological databases were used for meta-feature molecular characterization: Chaussabel et al.^17^ and Li et al.^18^ by means of the *tmod R package*^38^. The enrichment of genes in those modules was tested and functional terms below a hyper-geometric enrichment FDR < 0.01 were selected. The genes related to CpGs were obtained from the Illumina probe annotation.

### Healthy pattern characterization of SADs clusters

Healthy individuals were mapped to the clusters in order to define where in the model do the healthy molecular conditions fit. In this sense, the selected gene and CpG features were extracted from healthy individuals’ OMIC datasets and tested with the pre-defined model. Additionally, differential expression analysis was performed between the clusters and the healthy individuals. Transcriptome expression dataset samples and genes were filtered and cleaned as described before. Read count expression values were normalized using *voom* algorithm from the *limma R package*^30^. Known batch and potential confounders previously defined were included in the linear model. *limma package* was used for differential expression analysis. An FDR < 1e-5 and log2FC > |0.5| was the cut-off to consider transcripts as differentially expressed between healthy controls and SADs patients by cluster.

### Characterization for Clinical and Low-dimensional Data

The distribution across clusters of clinical annotations (diagnosis and clinical symptoms) and low-dimensional data (auto-antibodies, serum cytokine protein levels, cell subsets, and genetic risk markers) were analyzed. Numerical and categorical low-dimensional dataset associations with clusters were assessed using linear and logistic regression models, respectively. Features with Analysis of Variance (ANOVA) p-values below 0.01 were considered unevenly distributed across clusters, and z-scored beta coefficients were used to assess enrichments and depletions. *R packages car* ^39^ and *multcomp* ^40^ were used in the analysis. Genetic risk loci and clinical features were reduced to Principal Components (PCs) prior to regression analysis, only PCs with variance explained higher than 5 times the standard deviation plus the average of 1000 randomizations of the original matrix were tested (First 64 and 11 PCs were selected for risk associated SNPs and clinical features respectively, see Figure S5A and Figure S5B). Clinical features with less than 5% informative value (non-negative and non-missing values) were filtered before the analysis (Figure S5C) and the remaining missing values were imputed using *missMDA R package*^41^. Briefly, significant PCs were assessed by means of *estim_ncpPCA* function, and they were used for the imputation of the missing values using *imputePCA* function and regularized method. SADs risk alleles were obtained from: GWAS catalog and ImmunoBase databases. Only replicated risk alleles at genome-wide significance level (*p-value* < 5e10^−8^) for the main four SADs of the study (SLE, RA, pSjS and SSc) were downloaded and merged without redundancy.

### Inception cohort analysis

Inception cohort time points were assigned individually to clusters by means of the SNF model trained with the cross-sectional cohort. The results were analyzed by pair of samplings and samples with complete information: patients with information for baseline (M000) and 6 months (M006) samplings, n=103; M000 and 14 months (M014) samplings, n=78 and complete sampling points, n=68. The classification of patients for complete samples was as follows: *stable patients*, patients with the same cluster assignment for all the time points; *relapse – remission patients*, patients with assignments only to an aberrant cluster (inflammatory, lymphoid and interferon) and assignments to healthy-like individual cluster at any time point; *unstable patients*, patients with assignments to different aberrant clusters across time.

### Data availability

Data will be transferred to ELIXIR^42^ and made available upon request and in agreement with the privacy rules of the European Union. Additionally, relationship between gene expression and DNA methylation datasets can be explored at http://bioinfo.genyo.es/precisesadsdata/.

## Data Availability

Data will be transferred to ELIXIR and made available upon request and in agreement with the privacy rules of the European Union.

## ACKNOWLEDGMENTS

This work has been supported through a grant from the Innovative Medicines Initiative Joint Undertaking No. 115565 and in-kind and in-cash contributions from the EFPIA partners. G.B. is supported by the Instituto de Salud Carlos III (ISCIII, Spanish Health Ministry), through the Sara Borrell subprogram (CD18/00153). The authors would like to particularly express their gratitude to the patients, nurses and many others who helped directly or indirectly in the consecution of this study.

## AUTHOR CONTRIBUTIONS

**MEAR, LL, FMcD, EB, LB, J-OP** designed the study; **LL and LB** organized the electronic Clinical Report Form and supervised the clinical information; **RAQ, FJG, ALB, MRM, HNL**, prepared the protocols for biobanking, organized, registered, and processed the samples; **The PRECISESADS CLINICAL CONSORTIUM** agreed on the clinical data, recruited the patients, performed their clinical assessment, and obtained the samples and is composed by **IA, MB, AC, RF, FF, AM, AT, CV, ET, GM, BV, RBA, ACM, FG, MAGG, SR, BUG, RC, GE, IR-P, EDL, RL, DB, NH, NB, TW, ML, GS, MZ, MAA-Z, MCC-V, EC-E, AEC, MCFR, EdR, IDQ, YJG, IJM, NB, RO-C, NO, ER, CA, MG, PLM, TS, BL, A-LM, DC, VD-P, SJ-J, AS, CCh, DW, AB, MB, MD, SD, GK, LK, FH, ST**; **JMa and LL** monitored the patient recruitment, the informed consents and the ethical approvals; **LleL, CJ, CM, P-EJ, J-OP and the Flow Cytometry Consortium** composed by **DAE, TL, ET, NA, EN, NV, EDL, KK, NB, RL-P, AdG, JD, BR, QS, MA, AD, QC, VG, LK, AB, JC, MH-F** optimized and calibrated the flow cytometers across all recruitment centers, normalized, gated and procured the data; **GB, SB, ZM, ECM, MK, JRU, AGG, JK, AB, SH, JM, RL** performed RNASequencing, genotyping, methylation, data curation, and data processing; **MOB, JF, YR** performed the cytokine, small lipid moiety antibodies and coordinated the routine antibody laboratory tests; **JW** organized the data and prepared the data for analysis; **GB, RJB, CC, ERD, YI, LL, JM, FMcD, SR, MEAR, JF, JM, LB, J-OP, MH-F, MJ, TR, IW, EdR**, oversaw the progress of the study; **GB, FCM, RJB, YI, FMcD, MEAR, LB, CCh, MH-F, EB**, discussed and interpreted the results; **GB, SB, FCM, MMB, EC-M** performed the analyses; **JM-M, PCS, DT-D** produced the resource ADEx; **LL** compiled the clinical information and wrote the materials and methods section for the clinical data; and **GB** and **MEAR** wrote the entire manuscript. All authors approved of the content of the manuscript.

## AUTHOR INFORMATION

Correspondence should be directed to: Marta E. Alarcón-Riquelme, Department of Medical Genomics, Center for Genomics and Oncological Research (GENYO) Pfizer-University of Granada-Andalusian Regional Government. Health Sciences Technology Park, 18016, Granada, Spain.

